# Genomic landscape of endometrial, ovarian and cervical cancers in Japan from database in the Center for Cancer Genomics and Advanced Therapeutics

**DOI:** 10.1101/2023.10.09.23296710

**Authors:** Qian Xi, Hidenori Kage, Miho Ogawa, Asami Matsunaga, Akira Nishijima, Kenbun Sone, Kei Kawana, Katsutoshi Oda

## Abstract

**Objective:** This study aimed to comprehensively clarify genomic landscape, and its association with tumor mutation burden-high (TMB-H, ≧10 mut/Mb) and microsatellite instability-high (MSI-H) in endometrial, cervical, and ovarian cancers.

**Methods:** We obtained genomic datasets of a comprehensive genomic profiling test, FoundationOne® CDx, with clinical information by using the “Center for Cancer Genomics and Advanced Therapeutics” (C-CAT) database in Japan. The patients could take the tests only after the standardized treatments under universal health insurance coverage.

**Results:** Endometrial cancers (n=561) were characterized by high frequency of tumor mutational burden-high (TMB-H) (13.9%) and MSI-high (MSI-H) (10.8%), especially in endometrioid carcinomas. The lower ratio of *POLE* exonuclease mutations (1.4%) and higher ratio of *TP53* mutations (54.4%) than previous reports suggested the prognostic impact of the molecular subtypes. Among 839 cervical cancers, frequent mutations of *KRAS* (32.2%), *TP53* (29.4%), *PIK3CA* (22.2%), *STK11* (22.2%), *CDKN2A* (18.3%), and *ERBB2* (16.7%) were observed in adenocarcinomas, while the ratio of TMB-H was significantly higher in squamous cells carcinomas (20.6%). Among 1,606 ovarian cancers, genomic profiling of serous (n=784), clear cell (n=333), endometrioid (n=92), and mucinous carcinomas (n=91) was characterized. Pathogenic mutations in the *POLE* exonuclease domain were linked to high TMB (TMB >100 mut/Mb), and the mutation ratio was low in both cervical (0.0%) and ovarian cancer (0.19%).

**Conclusion:** The C-CAT database is useful to provide mutational landscape of each cancer type and each histological subtype. As the dataset is collected exclusively from patients after the standardized treatments, the information of “druggable” alterations highlights the unmet needs for drug development in major gynecological cancers.

## INTRODUCTION

Comprehensive genomic profiling (CGP) tests broadly explore treatments based on individual genomic information ^1^. Until June 2023, three CGP tests have been clinically applicable in Japan, including a tumor-only panel, FoundationOne® CDx (F1CDx) assay, a panel of liquid biopsy, FoundationOne Liquid® CDx assay, and a tumor/normal paired panel, OncoGuide^TM^ NCC Oncopanel System ^2,3^. All the genomic profiling data and clinical information are transferred to the Center for Cancer Genomics and Advanced Therapeutics (C-CAT) under written informed consent (agreement ratio of 99.7%), and the data are available for research use ^3^. As the CGP tests under the universal health insurance system in Japan are only applicable to patients who have (already or almost) finished standardized treatments, the dataset is composed of poor prognostic cancer patients in all the cancer types, except for cancers of unknown primary. Liquid biopsy is limited to patients whose tissue specimens are not available or inappropriate for CGP, and up to date, F1CDx has been broadly tested (>75%) in Japan. C-CAT database enables us to understand the mutational landscape, tumor mutational burden (TMB), and microsatellite instability (MSI) status in any type of solid tumors with poor prognosis ^3^.

Endometrial, cervical, and ovarian cancers are three major types of gynecological malignancies. In these three cancers, platinum-based chemotherapy is usually used, and CGP tests are anticipated to find novel treatment options. In endometrial cancers, genomic alterations are common in the phosphatidylinositol-3 kinase (PI3K) pathway (such as *PTEN*, *PIK3CA,* and *PIK3R1*) and in the receptor tyrosine kinase/RAS/beta-catenin pathway ^4,5^. Notably, four major molecular subtypes have been identified: (i) *POLE* ultramutated (in exonuclease domain), (ii) MSI-high (hypermutated), (iii) copy-number low (mainly endometrioid), and (iv) copy-number high (serous-like) ^4,6^. Immunohistochemistry of mismatch repair (MMR) genes and TP53 can alternatively be considered as MSI-high (MSI-H) and copy-number-high, respectively ^7^. In cervical cancers, genomic alterations of *PIK3CA* are the most common (26%), followed by *EP300* (11%) and *FBXW7* (11%) ^8^. In ovarian cancers, genomic alterations of *BRCA1*/*2* (both germline and somatic) and *TP53* are common in high-grade serous ovarian carcinomas ^9,10^. Genomic alterations of *ARID1A* and *PIK3CA* are detected at 30-60% in endometriosis-associated ovarian carcinomas, i.e., endometrioid and clear cell ovarian carcinomas ^11^. Genomic alterations of *KRAS* and *BRAF* in the MAPK pathway, as well as *TP53*, are common in mucinous ovarian carcinomas ^12^.

Both MSI-high and TMB-high (TMB-H; ≧10 mutations/megabase (mut/Mb)) are used as companion diagnostics for an immune checkpoint inhibitor (ICI), pembrolizumab, in solid tumors ^13,14^. In addition to these tumor-agnostic indications, lenvatinib (a multi-tyrosine kinase inhibitor) plus pembrolizumab was approved in Japan for advanced/recurrent endometrial cancer since December 2021, regardless of the MSI status ^15^. Recently, two phase III, randomized, clinical trials showed statistically significant benefits of overall survival and/or progression-free survival in primary advanced or recurrent endometrial cancer, regardless of the status of mismatch-repair deficiency (dMMR) ^16,17^. In advanced/recurrent cervical cancer, pembrolizumab in combination with platinum-based chemotherapy (with or without bevacizumab) was approved in Japan since September 2022, regardless of the PD-L1 combined positive score (CPS), based on the phase III KEYNOTE-826 trial with a statistically significant overall survival (OS) benefit ^18^. Cemiplimab monotherapy was also approved in recurrent cervical cancer as a second-line or later treatment in Japan since December 2022, regardless of PD-L1 status ^19^. However, the prognostic benefits of ICI-containing regimen are significantly larger by the presence of MSI-H and/or dMMR in endometrial and PD-L1 markers in cervical cancers. In ovarian cancers, TMB-H or MSI-H is still the only indication for pembrolizumab, although many ongoing clinical trials include ICIs ^20^.

In the present study, we focused on mutational landscape, TMB and MSI status in endometrial, cervical, and ovarian cancers, by using the C-CAT database of F1CDx in Japanese patients.

## MATERIALS AND METHODS

### 1. Patient samples of F1CDx from the C-CAT database

This Japanese cohort study included 561 endometrial cancers, 839 cervical cancers, and 1,606 ovarian cancers, which were analyzed by F1CDx under health insurance coverage. The data were obtained from the C-CAT database, organized by the National Cancer Center of Japan, which stores the CGP data tests ^3^. The CGP tests in Japan are limited only to solid cancer patients who have finished (or are expected to finish) standard treatments with unresectable diseases. Therefore, the patients enrolled were generally poor prognostic and resistant to platinum-based chemotherapies in all three gynecological cancers. We logged into the C-CAT system to collect 3,006 out of 25,504 patients’ F1CDx data in three gynecological cancers (between June 2019 and May 2022). The histological subtypes in each cancer were summarized in Sup. Table 1. In this study, pure sarcomas were not included in endometrial cancer, while 2 sarcomas and 63 non-epithelial tumors were included in cervical cancer and ovarian cancer, respectively. This study was ethically approved by our institutional ethics committee (#2021341G) and the Information Utilization Review Board of C-CAT (#CDU2022-026N).

### 2. F1CDx testing

F1CDx is a tumor-only panel using DNA isolated from formalin-fixed, paraffin-embedded (FFPE) tumor tissue specimens, which can detect substitutions, insertions and deletions (indels), copy number alterations (CNAs), and gene rearrangements in 324 genes, as well as genomic signatures including MSI and TMB ^21^. MSI status is reported as “cannot be determined” when the quality is insufficient. TMB by F1CDx is determined by counting all synonymous and non-synonymous variants except for hot spot genomic alterations and is considered TMB-High when reported as ≥ 10 mut/Mb. In our study, all genetic variants, including single nucleotide variants, copy number alterations, and gene fusions, were annotated as pathogenic or likely pathogenic based on the OncoKB, and/or ClinVar. MSI-H and TMB-H are tumor-agnostically approved as CDx for pembrolizumab in solid cancers in Japan. In this study, cases with “cannot be determined” for either TMB or MSI were excluded from the analysis (31 endometrial, 70 cervical, and 80 ovarian cancers).

### 3. Statistical analysis and graphic representations

Quantitative variables were analyzed using one-way ANOVA (when normality was assumed) and the Kruskal-Wallis H test (when normality could not be assumed) for comparisons among the three groups. Pearson’s correlation test was used for the correlation analysis between the two groups. All reported P values were two-tailed, and P<0.05 was considered significant unless otherwise specified. All the graphs, calculations, and statistical analyses were performed using GraphPad Prism software 9.3.0 and R 4.2.0 software. The collation and visual analysis of alteration data were implemented by the “ComplexHeatmap” package in R.

## RESULTS

### 1. Genomic alteration profiles across cancer types

We analyzed genomic alterations (pathogenic or likely pathogenic) in the F1CDx from the C-CAT database in 561 endometrial, 839 cervical, and 1,606 ovarian cancer samples. The mutational landscape of frequently mutated (pathogenic or likely pathogenic) genes (top 30) in each cancer type and in each histological subtype was summarized in Sup. Fig. 1, and Fig. 1, respectively (A: endometrial, B: cervical, and C: ovarian cancer).

**Figure 1.**
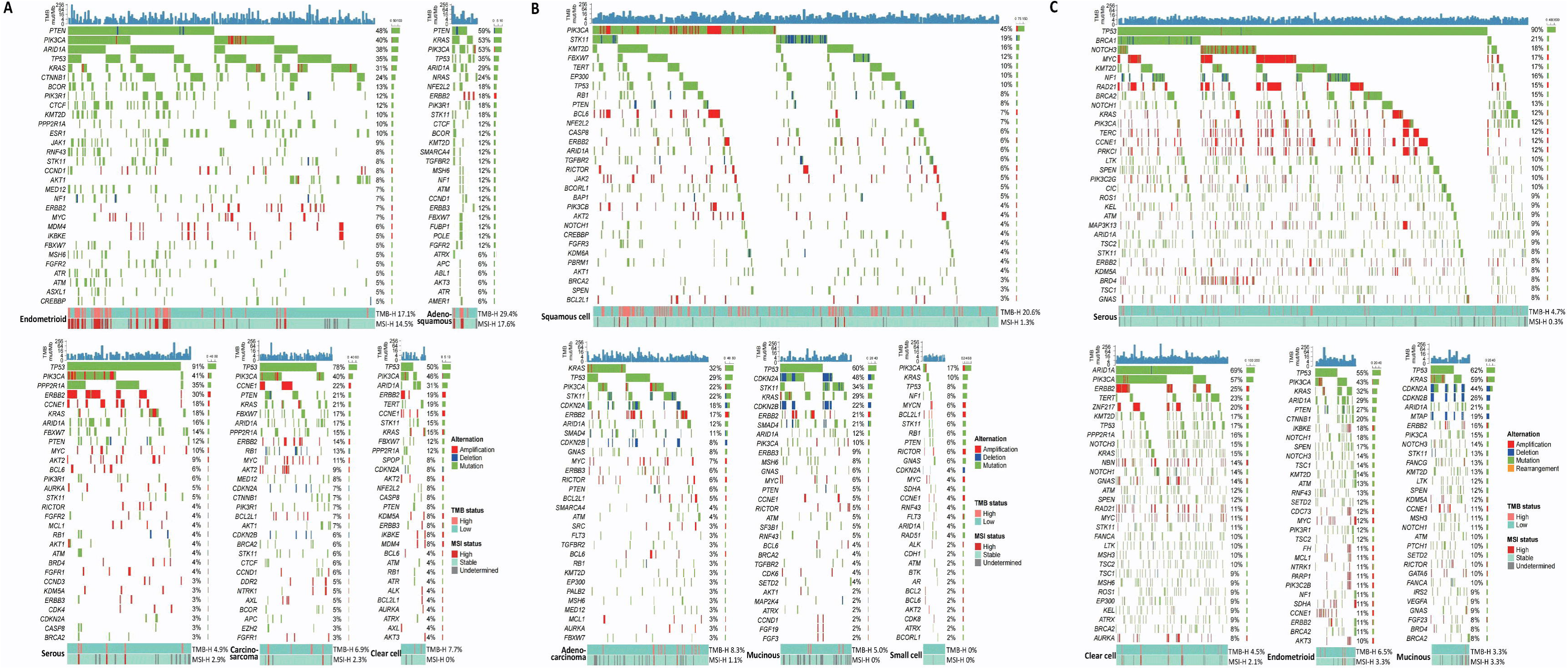
Genomic landscape of three gynecological cancers according to the histological subtypes. Recurrently mutated genes with types of alterations are listed with the status of TMB and MSI in (A) endometrial, (B) cervical, and (C) ovarian cancers. The upper plot represents the TMB scores by F1CDx.

#### Endometrial cancer

The five most frequent genomic alterations were *TP53* (n=305, 54.4%), *PIK3CA* (n=231, 41.2%), *PTEN* (n=194, 34.6%), *ARID1A* (n=172, 30.7%), and *KRAS* (n=146, 26.0%) (Sup Fig. 1A). The ratio of genomic alterations for these 5 genes in 232 endometrial cancer samples from TCGA dataset was 28.9% in *TP53*, 53.0% in *PIK3CA*, 63.8% in *PTEN*, 33.6% in *ARID1A* and 20.7% in *KRAS* ^9^. The ratio in *TP53* (P<0.0001) was significantly higher, and the ratio in *PTEN* (P<0.0001) and *PIK3CA* (P=0.0028) was significantly lower in the C-CAT database, compared with the TCGA database. In addition, the ratio of pathogenic/likely pathogenic alterations of *POLE* in exonuclease domain was only 1.4% (7.3% in TCGA), supporting the favorable prognosis of *POLE-*mutated endometrial carcinomas ^4^.

Endometrioid endometrial carcinoma, accounted for 49.0% of our study, was characterized by genomic alterations of *PTEN* (47.6% vs. 13.7%, P<0.0001), *KRAS* (30.9% vs. 17.8%, P=0.0037), *CTNNB1* (23.6% vs. 2.1%, P<0.0001), and *ARID1A* (37.8% vs. 22.6%, P=0.0015), compared with non-endometrioid endometrial carcinomas (serous, clear cell, and mixed carcinomas) (Fig. 1A). The high frequency of PI3K pathway aberrations at 69.8% (n=192) in endometrioid and *PIK3CA* genomic alterations at 43.8% (n=64) in non-endometrioid carcinomas suggested the requirement for potential targeted therapies in the PI3K pathway (Fig.2A). Genomic alterations of both *TP53* (80.8% vs. 35.3%, P<0.0001) and *ERBB2* (27.4% vs. 6.9%, P<0.0001) were more frequent in non-endometrioid carcinomas (Fig. 2A).

**Figure 2.**
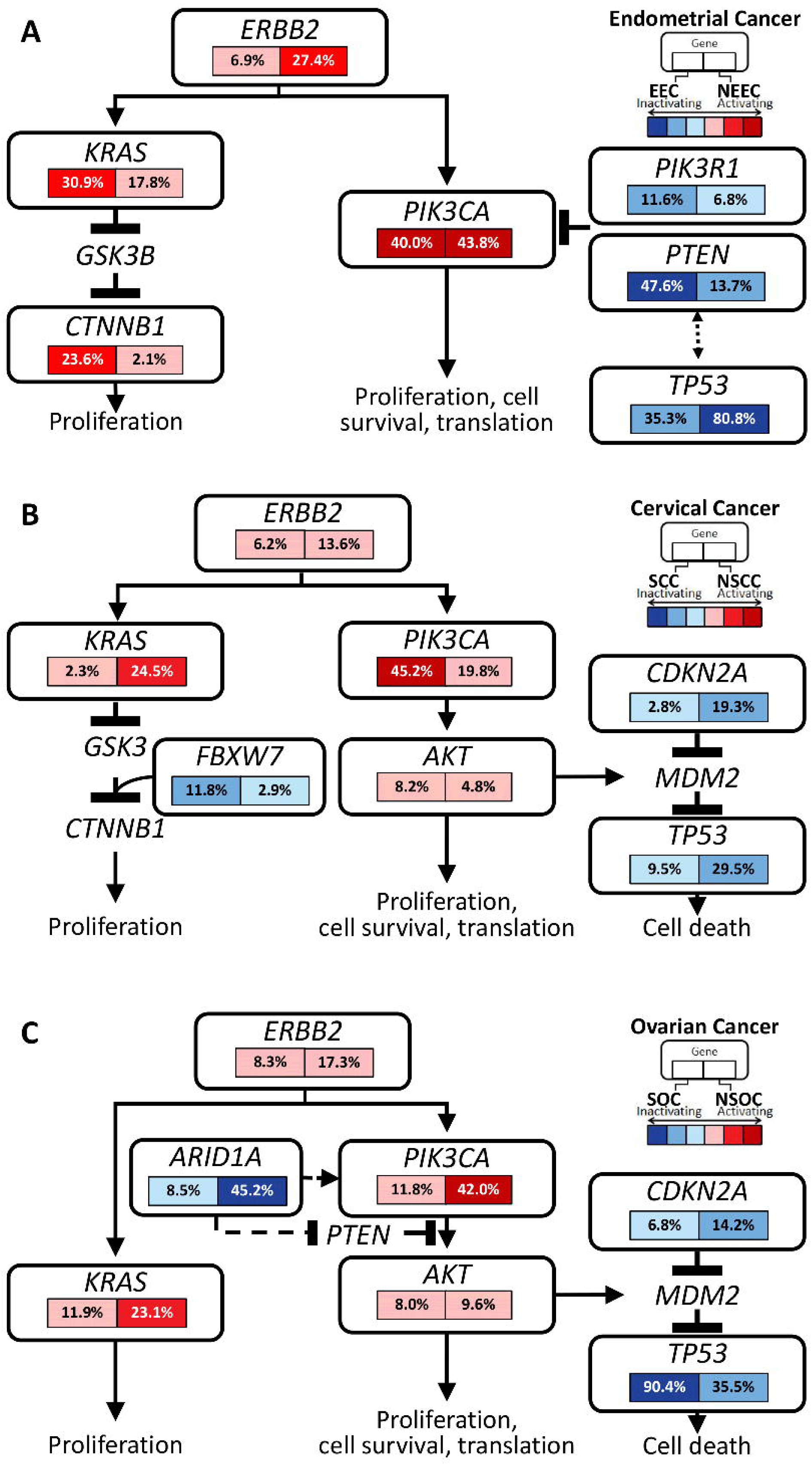
Frequency of genomic alterations in the key signaling pathways (mainly focusing on ERBB2, KRAS, and PI3K pathways) according to major histological types in (A) endometrial, (B) cervical, and (C) ovarian cancer. EEC: endometrioid endometrial carcinomas; NEEC, non-endometrioid endometrial carcinomas; SCC, squamous cell carcinomas; NSCC, non-squamous cell carcinomas; SOC, serous ovarian carcinomas; NSOC, non-serous ovarian carcinomas.

#### Cervical cancer

In the 839 cervical cancers, genomic alterations of *PIK3CA* were the most prevalent (n=270, 32.2%), followed by *STK11* (n=170, 20.3%), *TP53* (n=166, 19.8%), *KRAS* (n=117, 13.9%), and *CDKN2A* (n=96, 11.4%) (Sup. Fig. 1B). *ERBB2* genomic alterations were observed at 9.7% (amplifications at 6.3% and pathogenic variants at 4.1%), which might lead to clinical trials, including a “Basket Study of Tucatinib and Trastuzumab in Solid Tumors” (NCT04579380). Squamous cell carcinomas exhibit a significantly higher *PIK3CA* mutation rate of 45.2%, contrasting with 19.8% in non-squamous cell cases (Fig.2B). For *KRAS*, *CDKN2A*, and *TP53* in non-squamous cells, rates are 24.5%, 19.3%, and 29.5% respectively, while in squamous cell carcinoma rates are notably lower at 2.3%, 2.8%, and 9.5% (Fig.2B). In adenocarcinomas, *KRAS* genomic alterations were frequently observed at 32.2%, followed by *TP53* (29.4%), *PIK3CA* (22.2%), *STK11* (22.2%), *CDKN2A* (18.3%), *ERBB2* (16.7%), and *ARID1A* (11.7%) (Fig 1B). In mucinous carcinomas, the ratio of genomic alterations in *TP53* was 60.0%, followed by *CDKN2A* (47.5%), *STK11* (33.8%), and *KRAS* (28.8%), *CDKN2B* (22.5%), and *ERBB2* (21.3%).

#### Ovarian cancer

Among 1,606 ovarian cancers, *TP53* genomic alterations (n=1,054, 65.6%) were the most frequent, followed by *ARID1A* (n=407, 25.3%), *PIK3CA* (n=406, 25.3%), *KRAS* (n=272, 16.9%), *KMT2D* (n=272, 16.9%) and *NOTCH3* (n=270, 16.8%) (Sup Fig. 1C).

In serous carcinomas, genomic alterations of *BRCA1* and *BRCA2* accounted for 21.2% (166/784) and 14.7% (115/784), respectively (Fig. 1C). Overall, the ratio of genomic alterations in *BRCA1* and/or *BRCA2* was 32.7% (n=256). The coexistence rate of these two alterations was 4.8% (38/784), which was significantly higher than that reported by 0.6% (2/316) ^12^ and 0% (0/205) ^28^. Pathogenicity should be carefully interpreted, as the F1CDx and the C-CAT database annotation might be distinct from the other companion diagnostics (i.e. BRACAnalysis®). Genomic alterations in the other homologous recombination repair genes included *ATM* (8.8%), *PALB2* (7.1%), and *CDK12* (6.6%) (Fig. 1C). Genomic alterations in *TP53*, *NF1, KRAS*, and *PIK3CA* were detected at 90.4% (n=709), 15.8% (n=124), 11.9% (n=93), and 11.7% (n=92), respectively. Serous carcinomas were characterized by higher mutation rates for *TP53*, and lower mutation rates for *KRAS*, *PIK3CA*, and *ARID1A*, compared with non-serous ovarian cancers (Fig.2C).

Clear cell carcinomas were examined at 20.7% (n=333), with genomic alterations in *ARID1A* (n=231, 69.4%) and *PIK3CA* (n=190, 57.1%), compatible with previous reports ^15^ (Fig.1C). Genomic alterations of *TP53* were at 16.5% (n=55), and were negatively associated with alterations in both *ARID1A* (P<0.0001) and *PIK3CA* (P<0.0001) (Sup. Fig. 1C). Genomic alterations of *ERBB2* (mainly amplification) and *KRAS* were detected at 25% and 15%, respectively.

In endometrioid carcinomas, the ratio of genomic alterations in *TP53*, *PIK3CA*, *KRAS ARID1A*, *PTEN* and *CTNNB1* was 55.4%, 43.5%, 31.5%, 29.3%, 27.2%, and 19.6%, respectively. *TP53* alterations were negatively associated with alterations in *ARID1A* (P=0.0006), *KRAS* (P=0.0017), *PTEN* (P=0.0002), and *CTNNB1*(P<0.0001).

In mucinous carcinomas, genomic alterations of *TP53*, *KRAS*, *CDKN2A*, and *CDKN2B* were detected at 61.5%, 59.3%, 44.0%, and 26.4%, respectively. Although genomic alterations of *BRAF* were reported to be approximately 20% ^16^, the ratio was only 5.5% (n=5) in this study. Genomic alterations of *ERBB2* were detected at 16.5%.

### 2. Prevalence of MSI-H and TMB-H

Among 561 endometrial cancers, TMB-H and MSI-H were 78 (13.9%) and 61 (10.9%), respectively. Fifty-eight of 61 MSI-H tumors were TMB-H, while 20 of 78 (25.6%) with TMB-H were non-MSI-H (Fig. 3A). Among 839 cervical cancers, the number of TMB-H and MSI-H were 119 (14.2%) and 13 (1.5%), respectively (Fig. 3B). Only 1 of 13 (7.7%) cervical cancers with MSI-H was TMB-Low (TMB-L) (Fig. 3B). Among 1,606 ovarian cancers, the number of MSI-H and TMB-H was 80 (5.0%) and 19 (1.2%), respectively (Fig. 3C). All the 19 MSI-H ovarian cancers were TMB-H (Fig. 3C).

**Figure 3.**
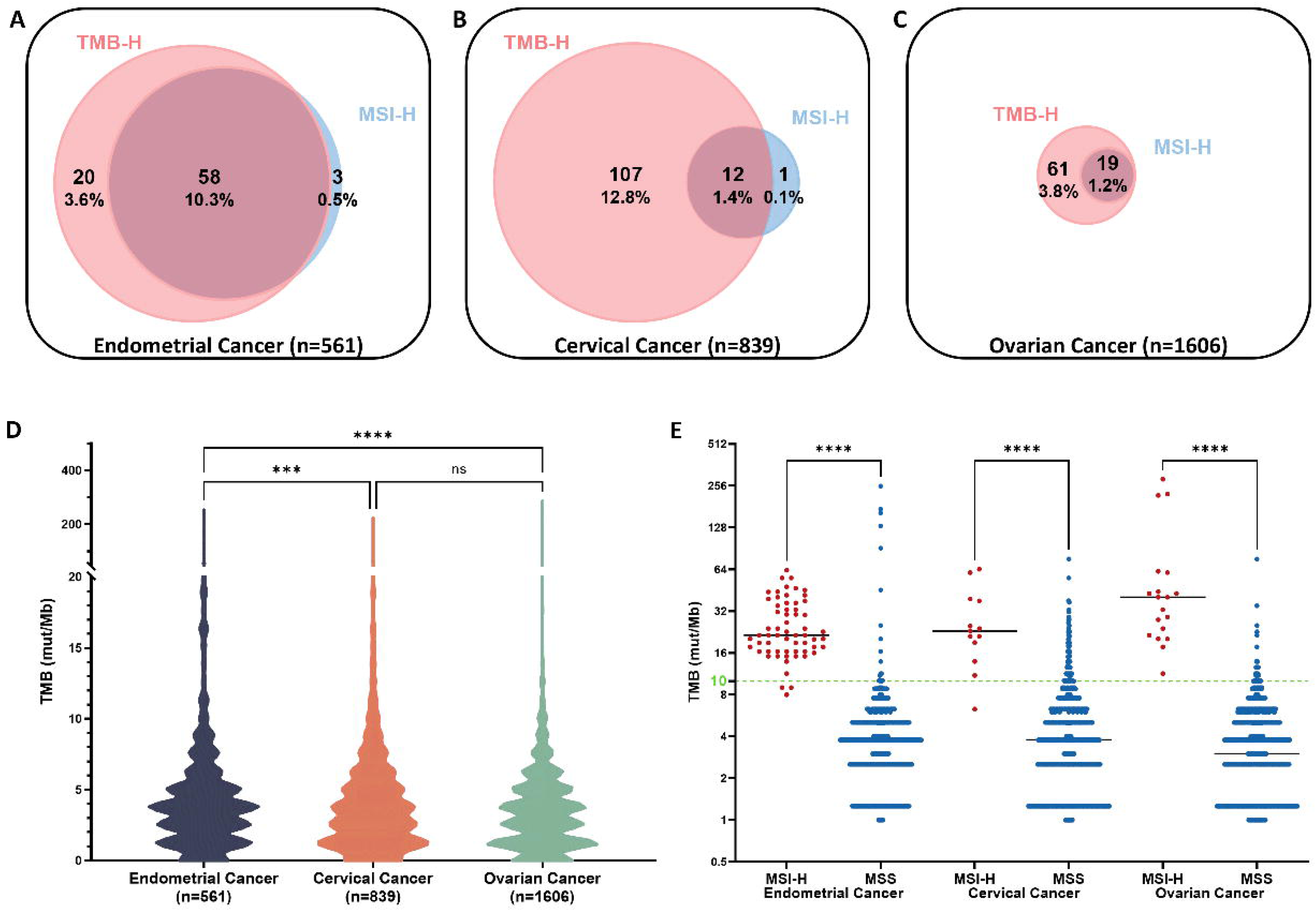
Venn diagrams of TMB-H and MSI-H in (A) endometrial, (B) cervical, and (C) ovarian cancer. (D) Violin plots of TMB levels, and (E) TMB distribution according to the MSI status in each cancer. *** P value < 0.001, and **** P value < 0.0001.

The value of TMB in endometrial cancer was significantly higher than that in cervical (P<0.001 by two-away ANOVA) and in ovarian cancer (P<0.0001) (Fig. 3D). The median TMB value in MSI-H tumors was 21.4 mut/Mb in endometrial, 23.0 mut/Mb in cervical, and 40.4 mut/Mb in ovarian cancer (Fig. 3E), with a strong correlation between MSI and TMB in these three cancer types (P<0.0001) (Fig. 3E).

The status of TMB and MSI was distinct among histological subtypes of each cancer (Table 1). In endometrial cancer, the ratio of TMB-H was high in adenosquamous carcinomas (5/17, 29.4%), mixed carcinomas (5/18, 27.8%), and endometrioid carcinomas (47/275, 17.1%), while the ratio was only 4.9-7.7% in serous, clear cell carcinomas and carcinosarcomas (Fig. 4A). In cervical cancer TMB-H was more common in squamous cell carcinomas (80/389, 20.6%), followed by neuroendocrine (5/35, 14.3%), and adenosquamous carcinomas (6/46, 13.0%). The ratio of TMB-H in adenocarcinomas (8.3%, 15/180) and mucinous carcinomas (5.0%, 4/80) was significantly lower than that in squamous cell carcinomas (P=0.0002, P=0.0004, respectively) (Fig 4A). In ovarian cancer, the ratio of TMB-H was 3.3-6.5% in all the histological subtypes.

**Figure 4.**
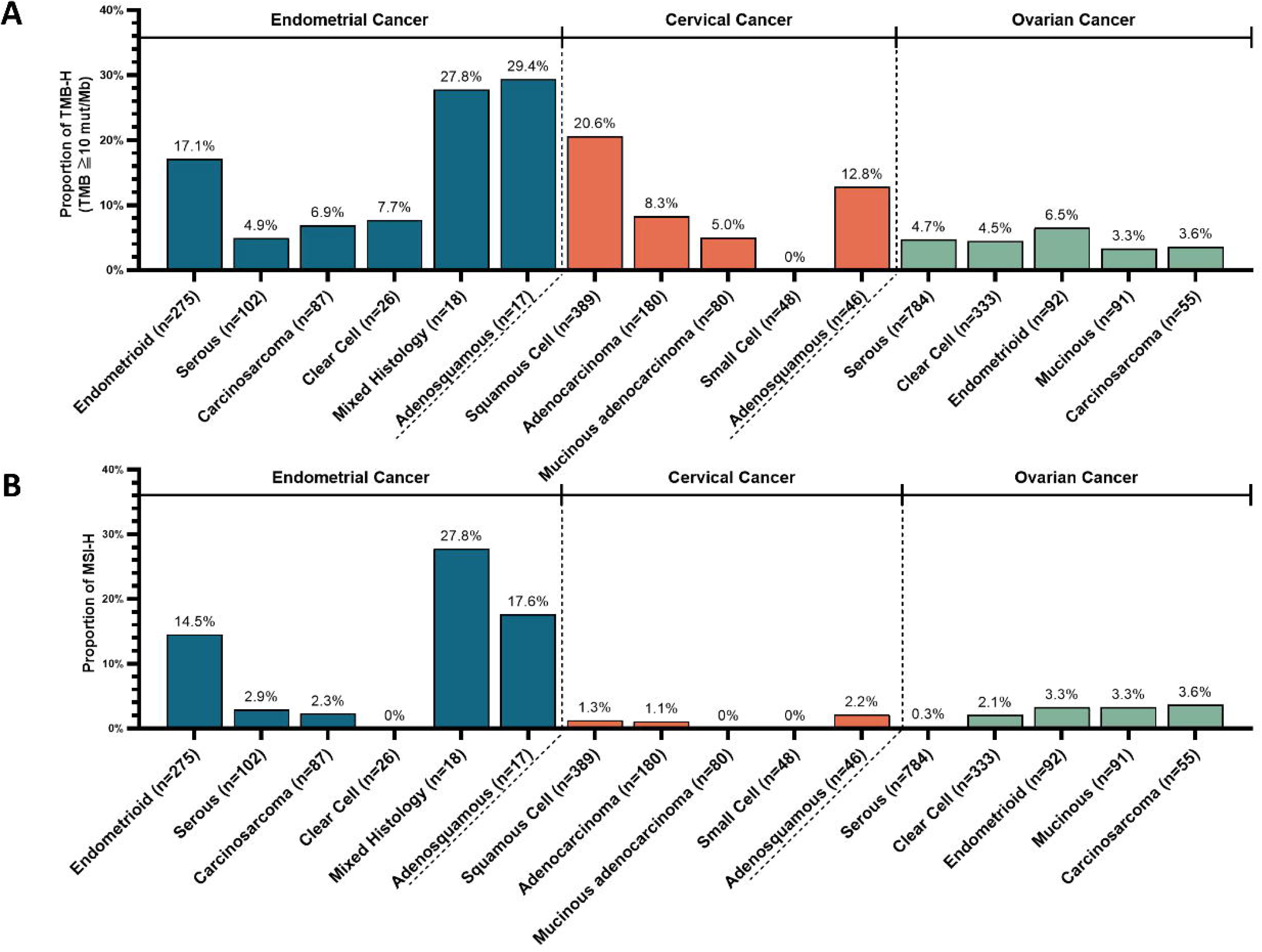
Frequency of (A) TMB-H and (B) MSI-H according to the major histological subtypes in endometrial, ovarian, and cervical cancer.

**Table 1.**
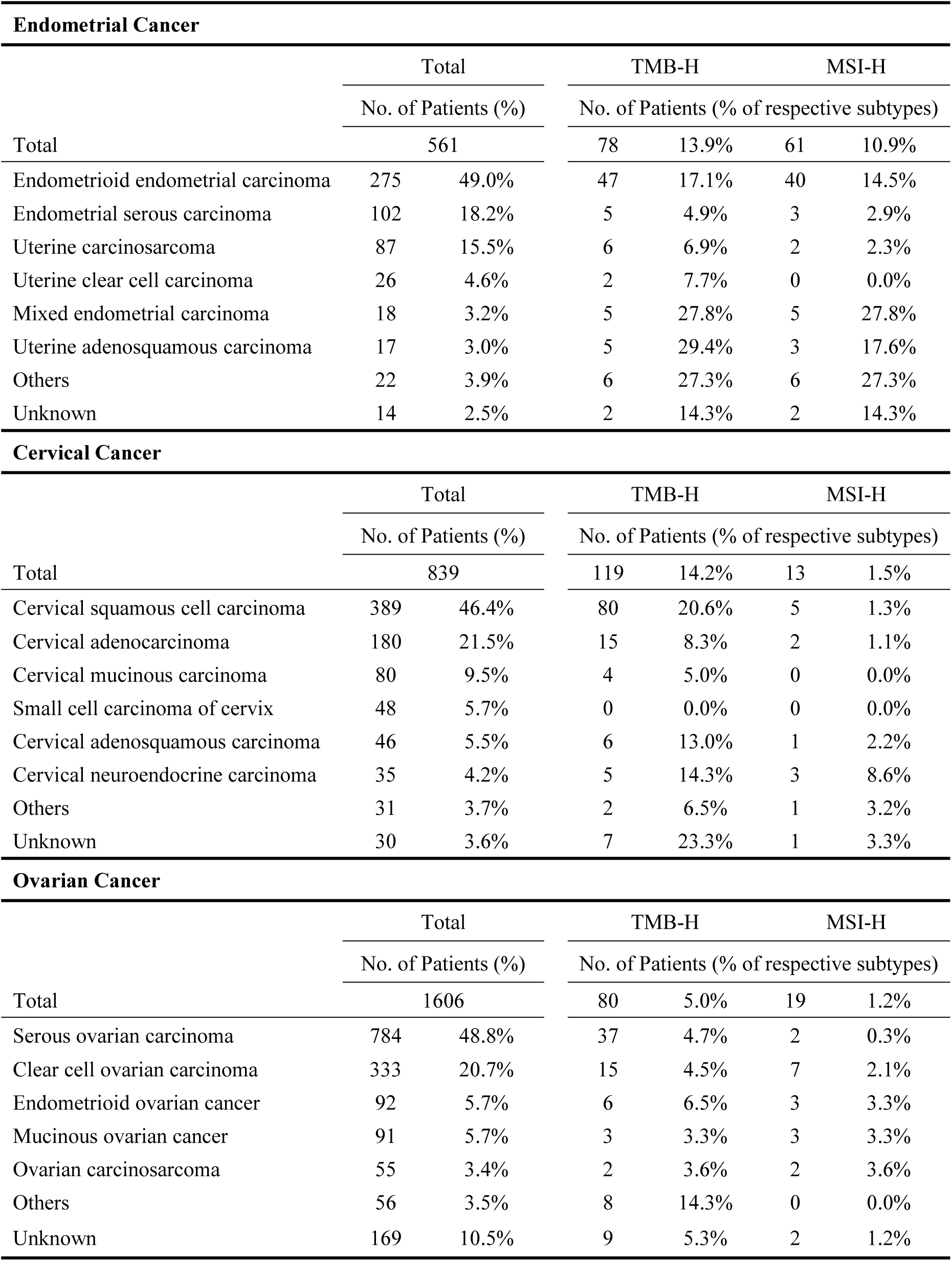
Frequency of TMB-H and MS-H according to the histological subtypes in each cancer.

Among various histological subtypes in endometrial cancer, the ratio of MSI-H was high in endometrioid (40/275, 14.5%), and adenosquamous carcinomas (3/17, 17.6%) (Table 1). The ratio of MSI-H in serous, clear cell, and carcinosarcomas (5/215, 2.3%) was significantly lower than that in endometrioid carcinomas (P<0.0001) (Fig. 4B). In cervical cancer, the ratio of MSI-H was not statistically significant between squamous cell carcinomas (1.3%) and adenocarcinomas (1.1%) (Fig. 4B). In ovarian cancer, the ratio of MSI-H was <4.0% in all the histological subtypes and significantly lower in serous carcinomas (2/784, 0.3%), compared with non-serous carcinomas (15/571, 2.6%), (P=0.0002) (Fig. 4B).

### 3. MMR-related mutations in tumors with MSI-H and TMB-H

We analyzed the correlation between genomic alterations of MMR genes (dMMR; defined as genomic alterations in *MLH1*, *PMS2*, *MSH2*, and *MSH6*) and MSI status in the three cancer types. In endometrial cancer, the ratio of dMMR was 31.1% (19/61) in MSI-H, which was significantly higher than 2.8% (13/469) in microsatellite stable (MSS) tumors (P<0.0001) (Sup Fig 2A). The ratio of dMMR in MSI-H and MSS in cervical cancer was 61.5% (8/13) and 4.6% (35/756) (P<0.0001), respectively, whereas that in ovarian cancer was 84.2% (16/19) and 13.5% (203/1,507) (P<0.0001), respectively (Sup. Fig. 2A).

Next, we analyzed the ratio of dMMR in MSS tumors and its association with TMB. The ratio of dMMR was significantly higher in TMB-H tumors (25%), compared with TMB-L tumors (2.5%) in MSS endometrial cancer (P=0.0003) (Sup. Fig. 2B). dMMR in MSS cervical cancer was also more frequent in TMB-H (9.4%), compared with TMB-L (4.3%) (P=0.0302). No statistical significance was detected in ovarian cancer (22.4% vs. 13.5%; P=0.0769) (Sup. Fig. 2B).

The highest prevalence of genomic alterations in MSI-H endometrial cancer was *MSH6* (n=14, 23.0%), followed by *MSH2* (n=8, 13.1%), *MLH1* (n=4, 6.6%), and *PMS2* (n=1, 1.6%) (Sup. Table 2). Similarly, such prevalence was also confirmed in ovarian cancer with MSI-H, with genomic alteration rates of *MSH6*, *MSH2*, *MLH1*, and *PMS2* of 52.6%, 36.8%, 31.6%, and 10.5%, respectively. In MSI-H cervical cancer, the genomic alteration rates of *MSH6* and *MLH1* were the highest (n=4,30.8%), followed by *MSH2* (n=2, 15.4%) and *PMS2* (n=1, 7.7%) (Sup. Table 2).

### 4. Distribution of *POLE* genomic alterations in exonuclease domain among TMB-H and MSS subsets

All the *POLE* variants (including variants of unknown significance: VUS) are listed in Table 2. Ultramutated genotype (TMB greater than 100 mut/Mb) was identified in 8 tumors (5 endometrial and 3 ovarian cancers). In endometrial cancer, all the 8 (1.4%) *POLE* exonuclease mutated tumors showed TMB-H (median TMB: 90.78 mut/Mb), only one of which was MSI-H (Table 2). Three MSI-H and TMB-H tumors showed VUS of *POLE* outside the exonuclease domain, which should be categorized as MSI-H, not as *POLE* subgroup (Table 2). Pathogenic/likely pathogenic variants in *POLE* exonuclease domain were detected at 1 case (0.12%) in cervical and at 3 (0.19%) in ovarian cancers. All the *POLE* variants outside the exonuclease domain were not annotated as pathogenic/likely pathogenic (Table 2).

**Table 2.**
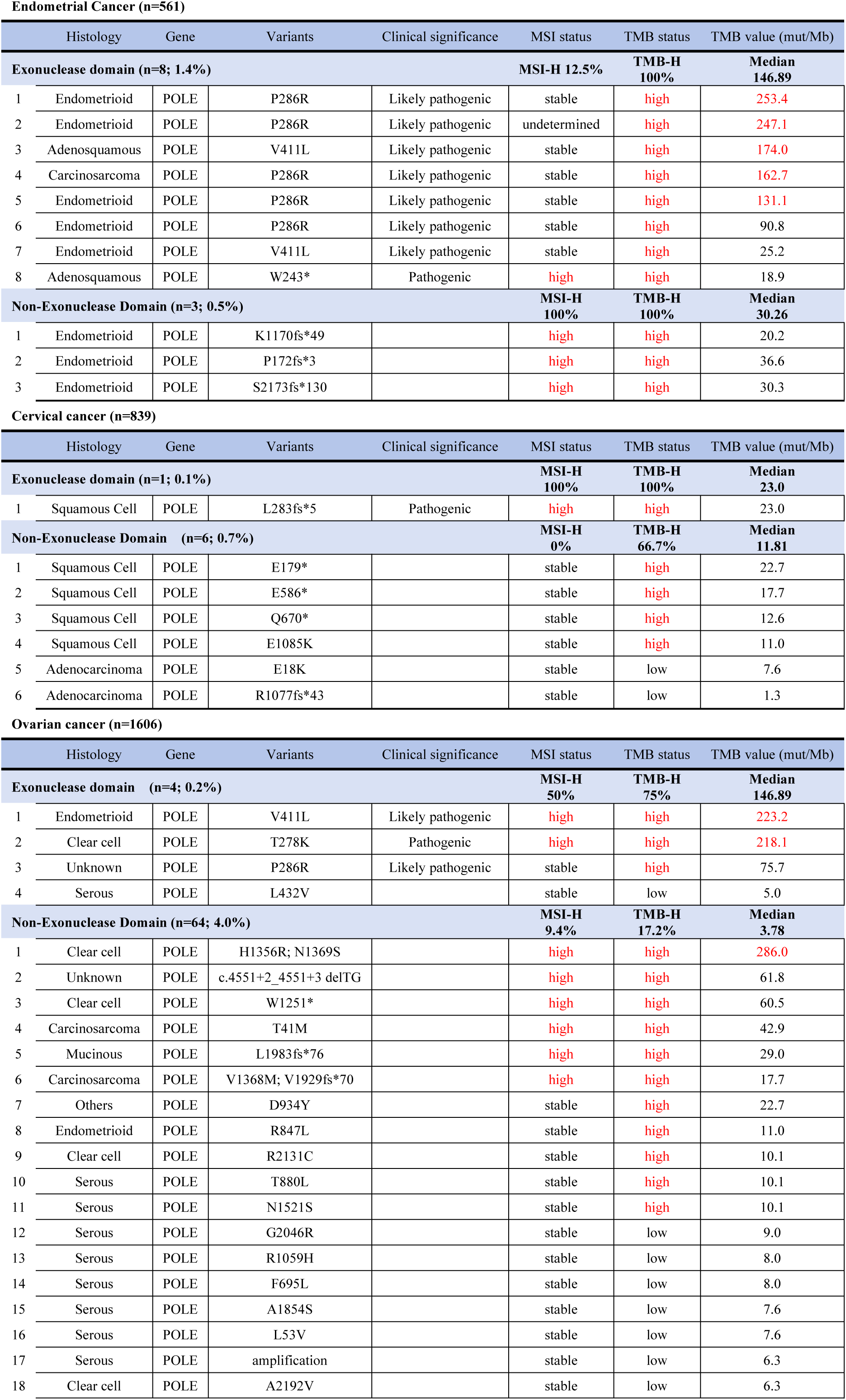
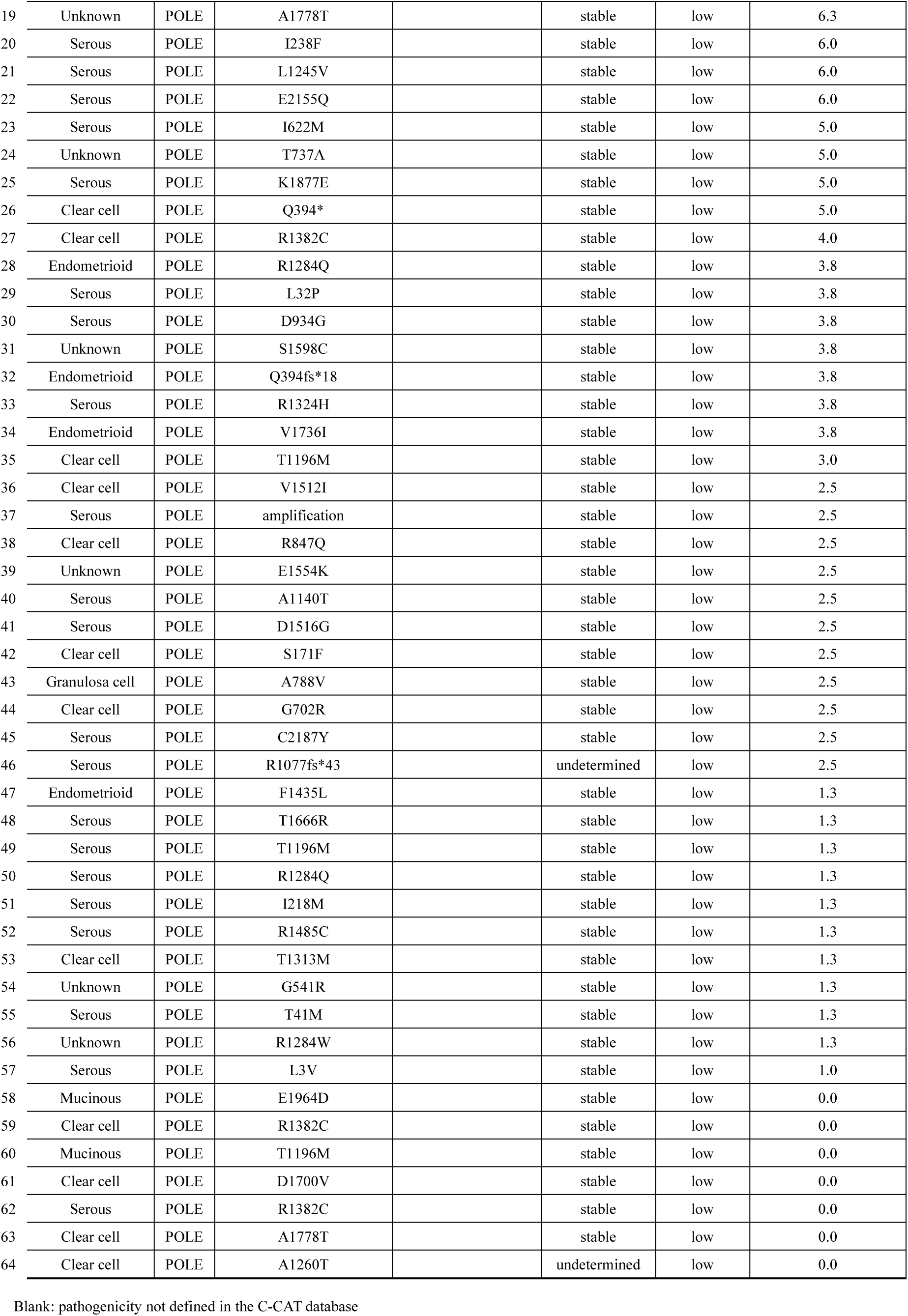
List of *POLE* variants with their pathogenicity, MSI and TMB status in each cancer.

### 5. Correlation among genomic alterations, MSI and TMB

Finally, we focused on the mutational landscape of “TMB-H with MSS” and “MSI-H” tumors in each cancer type. The most frequent genomic alteration in the “MSI-H” group was *ARID1A* in all three cancer types. The ratio was 96.7% (59/61) in endometrial, 76.9% (10/13) in cervical, and 89.5% (17/19) in ovarian cancer (Sup Fig. 3A-3C). Another MSI-H-related gene was *PTEN*. The ratio of genomic alterations of *PTEN* in the “MSI-H” group was 85.2% (52/61) in endometrial, 69.2% (9/13) in cervical, and 57.9% (11/19) in ovarian cancer, whereas the ratio of “MSS with TMB-L” was 28.2% (127/451) in endometrial, 7.5% (49/650) in cervical and 6.3% (92/1449) in ovarian cancer. The most frequent genomic alterations among MMR genes were *MSH6* in these three cancers with “MSI-H” (23.0% in endometrial, 30.8% in cervical, and 52.6% in ovarian cancer).

In “TMB-H with MSS” tumors, the ratio of genomic alterations in *PIK3CA* was the most or the 2^nd^ highest, which was 61.1% in endometrial, 51.4% in cervical, and 31.0% in ovarian cancer (Sup Fig. 3). Genomic alterations of *TP53* were the most common in the TMB-H with MSS group in endometrial (61.1%) and ovarian (82.8%) cancer, whereas the rate was 12.0% in cervical cancer (usually HPV-related with impairment of TP53 by ubiquitin-proteasome pathway). The ratio of genomic alterations in *CDKN2A,* and *CDKN2B* was also high in endometrial and ovarian cancer. The ratio was 16.7% for *CDKN2A,* and 16.7% for *CDKN2B* in endometrial cancer, whereas that was 22.4%, and 17.2% in ovarian cancer, respectively (Sup. Fig. 3).

## DISCUSSION

In this study, we analyzed totally 3,006 endometrial, cervical, and ovarian cancers with a tumor-only panel, F1CDx. The dataset of CGP tests in Japan is unique in eligible patients and insurance coverage. All the patients have finished or expected to finish the standardized treatments and taken the CGP tests under universal health insurance coverage ^3,22^. Thus, any poor-prognostic, Japanese cancer patients may have an opportunity to take the CGP tests. Furthermore, sufficient amount of tumor specimens is usually available through surgery and/or biopsy. Therefore, the C-CAT database is suitable for analyzing genomic profiling of poor prognostic patients in gynecological cancers.

In endometrial cancer, the comparison with the TCGA database highlighted the high incidence of genomic alterations of *TP53* (54.4%) and low incidence of genomic alterations of *POLE* (1.4%) in this database. This discrepancy supports the significance of the molecular classification of “Proactive Molecular Risk Classifier for Endometrial Cancer” (ProMisE) in endometrial cancer by POLE, dMMR, and TP53 ^23^. Indeed, ultramutated (>100 mutations/Mb) genotype in MSS was identified only in tumors with a *POLE* exonuclease mutation. The histology-based analysis highlighted the importance of the PI3K pathway (*PTEN* and *PIK3CA*), RAS pathway (*KRAS*), and wnt/β-catenin (*CTNNB1*) pathway in endometrioid carcinomas, and the significance of specific genes (*TP53*, *ERBB2* and *PIK3CA*) in non-endometrioid carcinomas. A WEE1 inhibitor, adavosertib, showed an ORR of 29.4% in recurrent uterine serous carcinomas (usually *TP53* mutated), and a phase IIb international study is ongoing ^24,25^. Further development of precision medicine in endometrial cancer is warranted.

In cervical cancer, the C-CAT dataset was helpful for elucidating genomic profiling of adenocarcinomas, as the ratio of non-squamous cell carcinoma was much lower in the TCGA dataset (19.1%) than that in the C-CAT database (53.6%) ^8^. Several druggable alterations were predominantly observed in adenocarcinomas, including *KRAS*, *ERBB2*, and *ARID1A.* According to the recently published 5th edition of the WHO Classification, cervical cancers are classified as HPV-associated and HPV-independent in each histological type ^26^. As both the TP53 and the RB pathway are impaired by HPV-E6 and E7 oncoproteins, respectively, genomic alterations of *TP53* and *RB* and *CDKN2A*/*2B* are informative to speculate HPV-independent cervical cancers, especially in gastric-type, mucinous adenocarcinomas ^27,28^.

In ovarian cancer, one limitation of the C-CAT database is that the data on low-grade serous ovarian carcinomas is mixed with high-grade serous carcinomas. The genomic alterations of *TP53* at 90% in serous carcinomas suggested that most are high-grade. The RAS-MAPK signaling pathway (genomic alterations of *NF1* at 16% and *KRAS* at 12% with mutual exclusivity), PI3K-mTOR pathway (*PIK3CA* at 12%, and *TSC2* at 8%), and certain receptor tyrosine kinases (*ROS1* at 9% and *ERBB2* at 8%) might be candidates for targeted therapy in serous carcinomas. As the ratio of genomic alterations of *BRCA1* (21.2%), *BRCA2* (14.7%), and its coexistence (4.8%) was higher than expected ^9,29^, the pathogenicity of each alteration should be carefully addressed. The high prevalence of genomic alterations of *ARID1A* and *PIK3CA* in clear cell ovarian carcinomas should be a key for novel therapies. Currently, a p110alpha selective inhibitor, CYH33, has been under phase 2 clinical trial (NCT05043922, jRCT2031210216), which recruits clear cell ovarian carcinoma patients with hot-spot mutations in *PIK3CA* ^30^. In endometrioid carcinomas, the inverse correlation between *TP53* and other genes (*PIK3CA*, *KRAS*, *ARID1A*, *PTEN,* and *CTNNB1*) suggested the importance of molecular profiling ^31^. In mucinous carcinomas, the high frequency of genomic alterations in *TP53*, *KRAS,* and *CDKN2A* is compatible with previous reports ^32^. The frequency of *BRAF* alterations has been reported to be from 5.4% to 22.6% ^32,33^. However, the ratio of *BRAF* alterations was 5.5% in this study, suggesting that *KRAS* is the leading driver of the RAS-MAPK pathway in mucinous carcinomas.

Candidate tumor-agnostic molecular targets in the 3 gynecological malignancies may include *ERBB2*, *PIK3CA*, *ARID1A*, and *KRAS*. An Antibody-Drug Conjugate, trastuzumab deruxtecan, showed an overall response rate of 54.5-70.0% in endometrial carcinosarcomas with positive HER2 in the STATICE trial ^34^. Genomic alterations of *ARID1A* may lead to novel molecular targeted therapies, including an EZH2 inhibitor and an enzyme for antioxidant glutathione synthesis ^35,36^. p110alpha selective inhibitors (alpelisib, etc), KRAS^G12C^ inhibitors (sotorasib, etc), KRAS^G12D^ degrader (ASP3082), a CBP/β-catenin inhibitor (E7386), which may be candidates ^37–40^. Japanese Gynecologic Oncology Group is currently conducting a basket trial of niraparib monotherapy for any gynecological cancers (except for ovarian cancer) with *BRCA1*/*2* genomic alterations, which targets the rare fraction of each cancer type ^41^.

MSI-H was reported to be the most frequent in endometrial cancer (16.9%) among 30 cancer types, and MSI-H in cervical and ovarian cancer accounted for 1.8% and 2.2%, respectively ^42^. The prevalence of TMB-H was reported at 25% in endometrial, and 17% in cervical cancers ^43^. Compatible with previous findings, MSI-H in this study was the main causative genomic finding to induce TMB-H in endometrial cancer, whereas it shared only 10% and 24% of the TMB-High in cervical and ovarian cancers, respectively. Low ratio of TMB-H (5.0%) in ovarian cancer may be associated with its limited sensitivity to ICI ^44^. We should be careful about passenger mutations in TMB-H tumors, as TMB-H, regardless of MSI status, may cause abundant genomic alterations. In this context, a comparison between “TMB-H with MSS” and “MSI-H” in each cancer type is informative to elucidate real “driver” alterations. In endometrial and ovarian cancers, the frequency of genomic alterations in *TP53*, *CDKN2A*/*2B* was significantly higher in the group of “TMB-H with MSS”. In cervical cancer, “TMB-H with MSS” was characterized by histology of squamous cell carcinomas and frequent genomic alterations of *PIK3CA*, *TERT,* and *STK11*, whereas “MSI-H” tumors were enriched with frequent alterations in *ARID1A* and *PTEN.* These findings suggest that TMB-H needs to be sub-classified according to the MSI status. Generally, genomic alterations in the MMR genes may be considered as MSI-H-like. However, we should pay attention that TMB-H with MSS tumors may contain genomic alterations of MMR genes, possibly as “passenger mutations”.

This study has several limitations. Firstly, the CGP tests in Japan are reimbursed only in patients who have (almost) finished standardized treatments, suggesting that patients with rapid progression might miss the opportunity to take the CGP tests. Secondly, response to genome-matched therapies has not been analyzed in this study, considering low accessibility to the recommended drugs. Thirdly, the C-CAT database is deposited from the designated hospitals located only in Japan. Therefore, most of the patients are Japanese.

In conclusion, genomic profiling of three major gynecological malignancies highlighted the mutational landscape of each cancer type, which is useful to clarify “druggable” alterations and the significance of molecular classification. Further, both TMB and MSI are essential to determine the tumor biology. The C-CAT database is useful to provide a comprehensive genomic profiling in the Japanese cohort, which may lead to future drug development.

## Data Availability

All data produced in the present study are available upon reasonable request to the authors.

## Author Contributions

Conceptualization and Validation: Q.X., H.K., M.O., A.M., A.N., K.S., K.K., K.O.

Data curation, Formal analysis, Investigation, Methodology, Project administration, Resources: Q.X., K.O.

Funding acquisition: K.O.

Software and Visualization: Q.X.

Supervision: H.K., M.O., K.S., K.O.

Writing – original draft: Q.X., K.O.

Writing – review & editing: Q.X., H.K., M.O., A.M., A.N., K.S., K.K.., K.O.

## Acknowledgements

This study was funded from the Grant-in-Aid for Scientific Research (B) (Grant number 21H03074 to K.O.). The text of the manuscript was edited by a native English speaker (https://www.editage.com).

