## Supplementary figures and images for "Genomic landscape of endometrial, ovarian and cervical cancers in Japan from database in the Center for Cancer Genomics and Advanced Therapeutics"

### Supplemental Figure 1

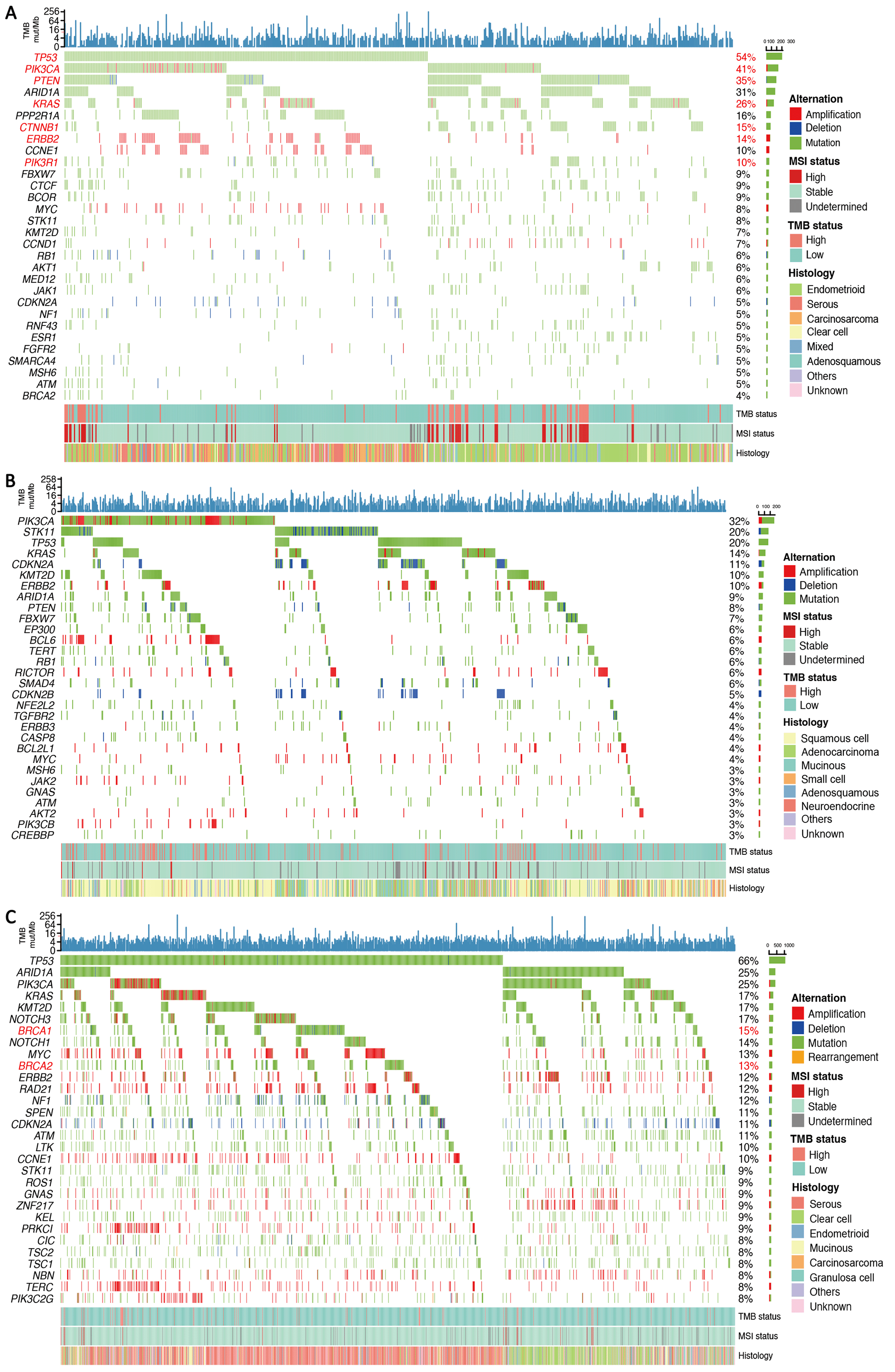

### Supplemental Figure 2

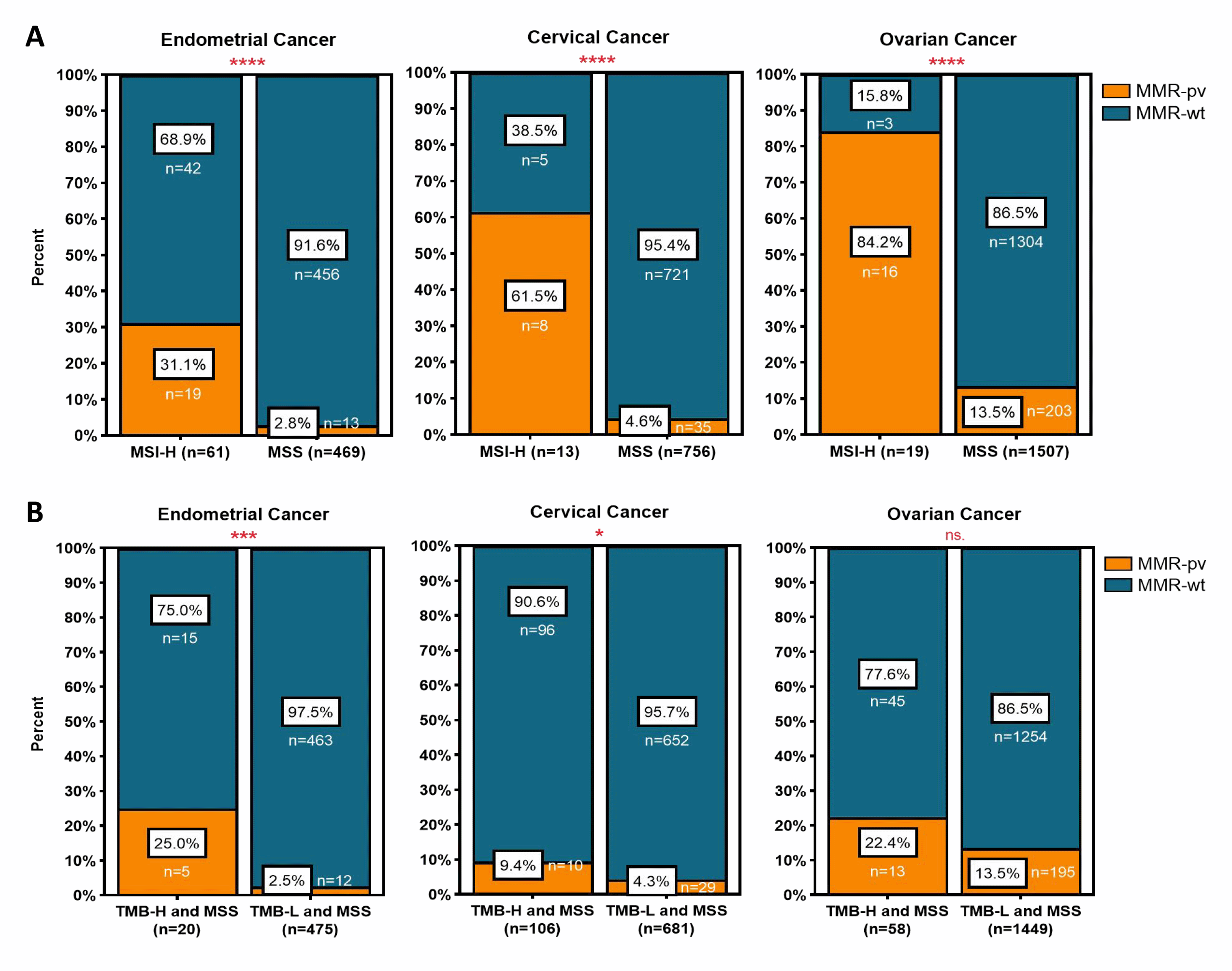

### Supplemental Figure 3

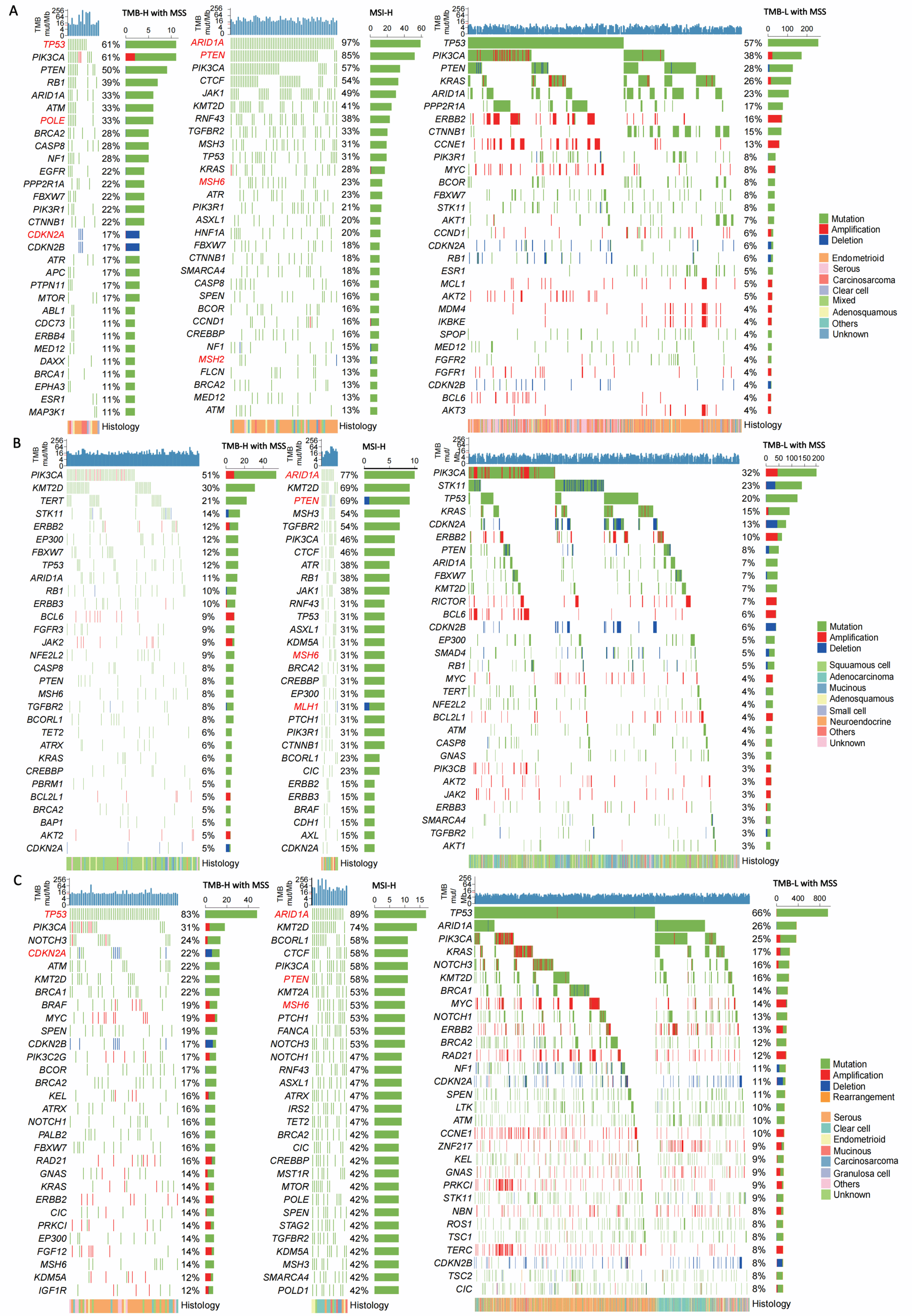

### Supplemental Table 1

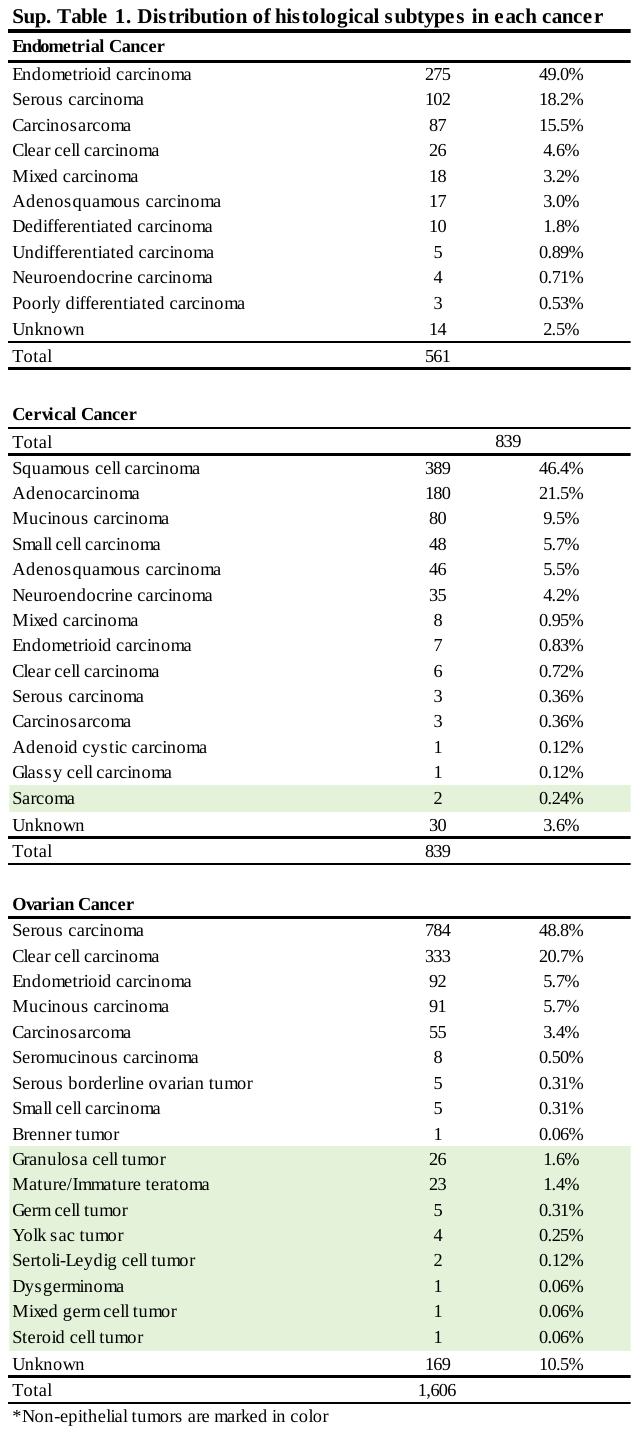
