## Supplemental Table 2 for "Genomic landscape of endometrial, ovarian and cervical cancers in Japan from database in the Center for Cancer Genomics and Advanced Therapeutics"

**Sup Table 2. Genomic alterations of MMR genes in each cancer with MSI-H**

|  | *MSH6* | | *MSH2* | | *MLH1* | | *PMS2* | |
| --- | --- | --- | --- | --- | --- | --- | --- | --- |
| Endometrial Cancer with MSI-H (n=61) | 14 | 23.0% | 8 | 13.1% | 4 | 6.6% | 1 | 1.6% |
| Cervical Cancer with MSI-H (n=13) | 4 | 30.8% | 2 | 15.4% | 4 | 30.8% | 1 | 7.7% |
| Ovarian Cancer with MSI-H (n=19) | 10 | 52.6% | 7 | 36.8% | 6 | 31.6% | 2 | 10.5% |
