## Supplementary Figure Legends for "Genomic landscape of endometrial, ovarian and cervical cancers in Japan from database in the Center for Cancer Genomics and Advanced Therapeutics"

**Supplementary Figure 1.** Genomic landscape of three gynecological cancers. Recurrently mutated genes are listed with the status of TMB and MSI and with information about types of alterations and histological subtypes in (A) endometrial, (B) cervical, and (C) ovarian cancers. The upper plot represents the TMB scores by F1CDx.Waterfall plot of genetic alteration profiles in endometrial (A), cervical (B), and ovarian cancer (C).

**Supplementary Figure 2.** Frequency of genomic alterations in the mismatch repair (MMR) genes according to the MSI and TMB stauts in each cancer type. (A) Frequency of MMR alterations according to the MSI status, (B) Frequency of MMR alterations according to the TMB status. Comparisons between the groups were performed by Fisher’s exact test (* P<0.05; ** P<0.01; *** P<0.001; ****P<0.0001). MMR-pv, pathogenic variants in MMR genes, MMR-wt, no pathogenic variants (wild-type) in MMR genes.

**Supplementary Figure 3.** Genomic landscape according to the TMB and MSI status in (A) endometrial, (B) cervical, and (C) ovarian cancer. Each cancer was categorized as TMB-H with MSS, MSI-H (regardless of TMB status), and TMB-L with MSS.
